# Retinal microvascular features are associated with CMR measures of subclinical cardiovascular dysfunction

**DOI:** 10.64898/2026.03.26.26348318

**Authors:** Cian Wade, Alicja R Rudnicka, Hana El Diwany, Charlotte Zheng, Ian Yeung, Robin D Hamilton, Masliza Mahmod, Helena B Thomaides Brears, Charlie Diamond, Ravi Pattanshetty, John Anderson, Ryan Chambers, Roshan A Welikala, Jiri Fajtl, Sarah A Barman, Elijah R Behr, Christopher G Owen

## Abstract

**Aims:** Microvascular dysfunction is implicated in the pathogenesis of many cardiovascular (CV) diseases, with retinal imaging providing a non-invasive window into microvascular health. Prior evidence links retinal microvascular features (RVF) with cardiac structure and function, yet these relationships remain incompletely characterised. This study systematically examined associations between RVFs and measures of subclinical CV dysfunction derived from CMR imaging.

**Methods and results:** 182 participants with type 2 diabetes were considered for inclusion in this cross-sectional study. Fifteen CMR measures of cardiac structure, function, tissue characterisation, adiposity, and aortic distensibility were derived. 128 participants (70%) had eligible retinal images. An artificial intelligence (AI)-enabled retinal image analysis tool (QUARTZ) quantified eight RVFs: arteriolar and venular diameter, area, calibre uniformity, and tortuosity. Multivariable regression examined RVF-CMR associations, expressed as change in CMR measures per 1-standard deviation (SD) increase in RVF. Five RVF-CMR associations remained significant after adjustment for confounders. Venular tortuosity was associated with a 0.5ms greater left ventricular (LV) T2 mean, 0.6% worsening in LV global longitudinal strain, and a 2 mL greater left atrial max volume. Arteriolar calibre uniformity and venular area were each associated with 9ms lower LV T1 mean and 0.2×10^-3^mmHg^-1^ greater proximal descending aortic distensibility, respectively.

**Conclusion:** In a diabetic cohort, we identified novel and biologically coherent associations between RVF and CMR measures. Notably, venular tortuosity was associated with a constellation of CMR changes consistent with subclinical myocardial dysfunction. These findings support the potential role of retinal imaging in evaluating CV dysfunction prior to overt disease.

## Introduction

Features of the retinal microvasculature are predictive of a range of cardiovascular (CV) diseases^1,2^. Numerous prospective studies have demonstrated an association between narrower retinal arteriolar diameters and coronary heart disease, hypertension, and heart failure^2,3^, while wider retinal venular diameter is associated with incident stroke^4^.

Beyond associations with clinically apparent, or overt, CV disease, evidence is emerging of the relationship between retinal microvascular features (RVFs) and indicators of subclinical CV dysfunction. Changes in arteriolar geometric features, including vessel branching angles and tortuosity are related to echocardiographic features of maladaptive cardiac remodelling^5^. Early features of macrovascular disease such as arterial and aortic stiffness are associated with narrower retinal arteriolar diameter^6,7^. Cardiovascular magnetic resonance (CMR) imaging is the gold-standard technique for characterisation of many subclinical CV abnormalities. A recent study examining RVFs against a limited set of CMR phenotypes found a significant correlation between central retinal venular equivalent and LV mass/volume ratio^8^. Notably however, when employing end-to-end deep-learning models, they found that retinal images had poor predictive performance for classifying binary outcomes of CMR-derived subclinical CV disease. This points to a clear evidence gap regarding the relationship between more granular measures of the retinal microvasculature and comprehensive CMR phenotypes.

CMR is not a point of care test and patients with subclinical disease will not routinely be referred for CMR, limiting its utility for diagnosing and monitoring early CV disease. The comparative accessibility and low-cost of retinal imaging has heightened interest in its utility as a non-invasive and scalable method of screening for CV disease. The case for using retinal imaging in this way rests on the retinal microvasculature providing a window into systemic vascular health^9^. Microvascular dysfunction is a hallmark of many CV diseases, frequently preceding and occurring in parallel to macrovascular disease due to both vascular beds’ shared susceptibility to systemic biochemical and haemodynamic factors^10,11,12^.

In this cross-sectional study, we systematically examined associations between features of the retinal microvasculature and CMR measures of subclinical CV dysfunction in a cohort of adults with type 2 diabetes. A key methodological advance of this study, compared to prior work, is the use of a fully-automated artificial intelligence (AI)-enabled system to quantify granular retinal microvascular features derived from the whole retinal image and examine these outputs against comprehensive CMR phenotypes.

## Methods

### Study design

The UK Imaging Diabetes Study (UKIDS, clinical trial registration number: NCT05057403) is an ongoing real-world, multi-centre prospective cohort study with clinical outcome collection of individuals with type 2 diabetes and without a recent history of incident CV disease. UKIDS participants recruited from Moorfields Eye Hospital NHS Foundation Trust in London, UK, between June 2022 – September 2025 were considered for inclusion. All participants had type 2 diabetes and were having regular outpatient retinal monitoring as part of their routine diabetic management. Exclusion criteria were evidence of incident CV event(s) within 12 months prior to consent, including myocardial infarction, unstable angina, heart surgery, ischaemic stroke, and transient ischaemic attack. Other exclusion criteria included known liver disease, known significant renal tract abnormalities, alcohol dependency, and contraindication to MR scanning.

### CMR data collection & processing

All participants underwent a standardised multi-organ multiparametric MR scan (CoverScan™; Perspectum, Oxford) on a 3T scanner at the Alliance Medical Imaging Centre, London, UK^13^. Acquired CMR images were processed by trained radiographers and MR technologists blinded to clinical data. Comprehensive CMR measures were derived, including: left ventricle ejection fraction (LVEF); LV end diastolic volume indexed to body surface area (BSA) (LVEDVi); LV end systolic volume BSA (LVESVi); LV stroke volume BSA (LVSVi), LA max volume BSA (LAVi) LV mass BSA (LVMi), LV wall thickness (LVWT), LV T1 and T2 mean relaxation times averaged across all 16 American Heart Association (AHA) myocardial segments derived from the three short-axis views excluding the apex, LV global longitudinal strain (LV GLS), LV global circumferential strain (LV GCS), LV global radial strain (LV GRS), epicardial adipose tissue (EAT), pericardial adipose tissue (PAT), and aortic distensibility measures (supplementary table 1). Data were centrally curated and quality-controlled. A cardiologist accredited to European Association of Cardiovascular Imaging level 3 for CMR independently reviewed all cardiac scans. Where pathology was identified, consensus was sought from a second independent cardiologist with this same accreditation. Scans were reviewed to exclude regional wall motion abnormalities, clinically significant valvular abnormalities, and other cardiac pathology. If a given CMR measure had a reporting completion of <70% across included participants, then this measure was not included in the analysis. Demographic and self-reported clinical information were collected from participants by UKIDS researchers during a face-to-face consultation prior to CMR images acquisition (supplementary table 2).

### Retinal image collection & processing

Macular centred retinal images of participants were searched for on imaging databases held at Moorfields Eye Hospital NHS Foundation Trust and the North East London Diabetic Eye Screening programme with the following selection criteria applied: colour fundus image of either eye captured with a 45^D^ field of view; retinal image captured within a 2-year period either before or after the date the CMR images were acquired; quality of retinal image ≥70% (as per QUARTZ methodology outlined below). Where multiple retinal images met the inclusion criteria, the single image with the highest quality was selected.

A fully-automated artificial intelligence (AI)-enabled system (QUARTZ)^14^ was applied to extract and quantify thousands of measures related to the retinal microvasculature within each single retinal image. QUARTZ uses deep learning to classify retinal vessels as an arteriole or venule, map the optic disc, and calculate an image quality score ranging from 0-100%. For this study, eight RVF measures were used from QUARTZ’s output: arteriolar and venular diameter (µm), area (mm^2^), calibre uniformity (1/µm), and tortuosity (arbitrary units). We define vessel calibre uniformity as the variability of the diameter of vessels within an individual’s retina. This is calculated by the median standard deviation of vessel diameter along vessel segments, computed per image and inverted to normalise. Therefore, greater vessel calibre uniformity indicates lower variability of diameter, and vice-versa. Pixels from each retinal image were converted to physical units (in microns) using the QUARTZ output of the optic disc vertical diameter as a calibration reference standard^15^. RVF diameter, calibre uniformity, and tortuosity were calculated for each identified vessel and an average (weighted by length of the vessel segment) calculated for each participant’s image. The RVF area measure represents the total arteriolar or venular area in each image.

### Statistics

Statistical analysis was carried out using Stata (StataCorp LLC. Stata Statistical Software: Release 19.5. College Station (TX); 2024). See supplementary table 3 for additional details on statistical methods. Participant demographic and clinical characteristics were reported, as well as RVF and CMR measure mean and standard deviation (where normally distributed) or median and interquartile range (where non-normally distributed). A participant was excluded from further analysis if any RVF or CMR measure had a |z| score >4, to minimise the influence of potentially artefactual retinal vessel or CMR measurements.

Pearson’s correlation was performed on all 8 RVFs paired with all CMR measures that met ≥70% reporting completion, excluding unindexed volumetric measures, yielding 120 RVF-CMR pairs. For each of the RVF-CMR pairs, participants with missing data for the CMR measure were excluded from that correlation analysis. To address multiple testing, we applied a multi-step selection strategy to shortlist RVF-CMR pairs for further regression analysis. First, we required associations to demonstrate both a minimum effect size (|r| ≥0.15) and statistical significance (p<0.05). Second, we evaluated the biological feasibility of relationships meeting this threshold.

Where normality violations were identified, log transformation was attempted and robust standard errors applied where violations persisted. Systolic blood pressure (SBP) was unrecorded among 16 (12.5%) participants. Following a Missing at Random (MAR) assessment, multiple imputation by chained equations with predictive mean matching was used to handle missing SBP data.

Multivariable linear regression was performed on multiply imputed datasets to predict CMR measures from RVF. Where a CMR measure was missing for the measure under analysis, participants were dropped without imputation. Sequential models 1-6 adjusted for age, sex, SBP, diabetes duration, and time between retinal and CMR image acquisition as potential confounders. Model 6 additionally incorporated a centre-specific fixed effect (α□) to account for potential variation in retinal image acquisition between sites (supplementary table 4).

Regression coefficients are reported as unstandardized (β represents change in CMR measure units per 1-standard deviation [SD] increase in RVF) and standardized (βSD represents change in SD units of CMR measure per 1-SD increase in RVF), with 95% confidence intervals and significance set at α=0.05. Sensitivity analysis assessed stability of β estimates to a ∼10% reduction in sample size (supplementary table 3e).

### Ethics

Informed written consent was obtained from all study participants enrolling in the UKIDS trial. Ethical approval for UKIDS was provided by the West Midlands – Solihull research ethics committee (ethics reference 20/WM/0007). Retinal imaging data used from the North East London Diabetic Eye Screening programme was approved by NHS Health Research Authority and Health and Care Research Wales (Integrated Research Application System number 265637). This study was conducted in accordance with the Declaration of Helsinki.

## Results

### Demographic & clinical characteristics

182 participants in the UKIDS study recruited from Moorfields Eye Hospital NHS Foundation Trust underwent CMR imaging between June 2022 and September 2025. 129 (71%) of these had retinal images which met the inclusion criteria. One participant (0.8%) was excluded due to extreme RVF measurements (venular diameter Z=4.2, arteriolar area Z=7.8, venular area Z=8.4), indicating likely imaging artefact. Therefore, 128 participants (70%) were included for analysis (Figure 1). No systematic demographic, RVF or CMR differences were observed between the 128 included and 54 excluded participants. Of the included participants, 88 (69%) were male, 40 (31%) were female and their mean age was 62 years (SD 9) (Table 1). 37% were of White ethnic origin, 32% South Asian, 23% Black, and 8% from other groups. Mean duration of type 2 diabetes was 17 years (SD 9), with mean SBP of 138mmHg (SD 17), diastolic blood pressure of 79mmHg (SD 9), and mean BMI of 29 kg/m² (SD 5). 8% were current smokers, 22% past smokers, and 70% had never smoked.

**Figure 1:**
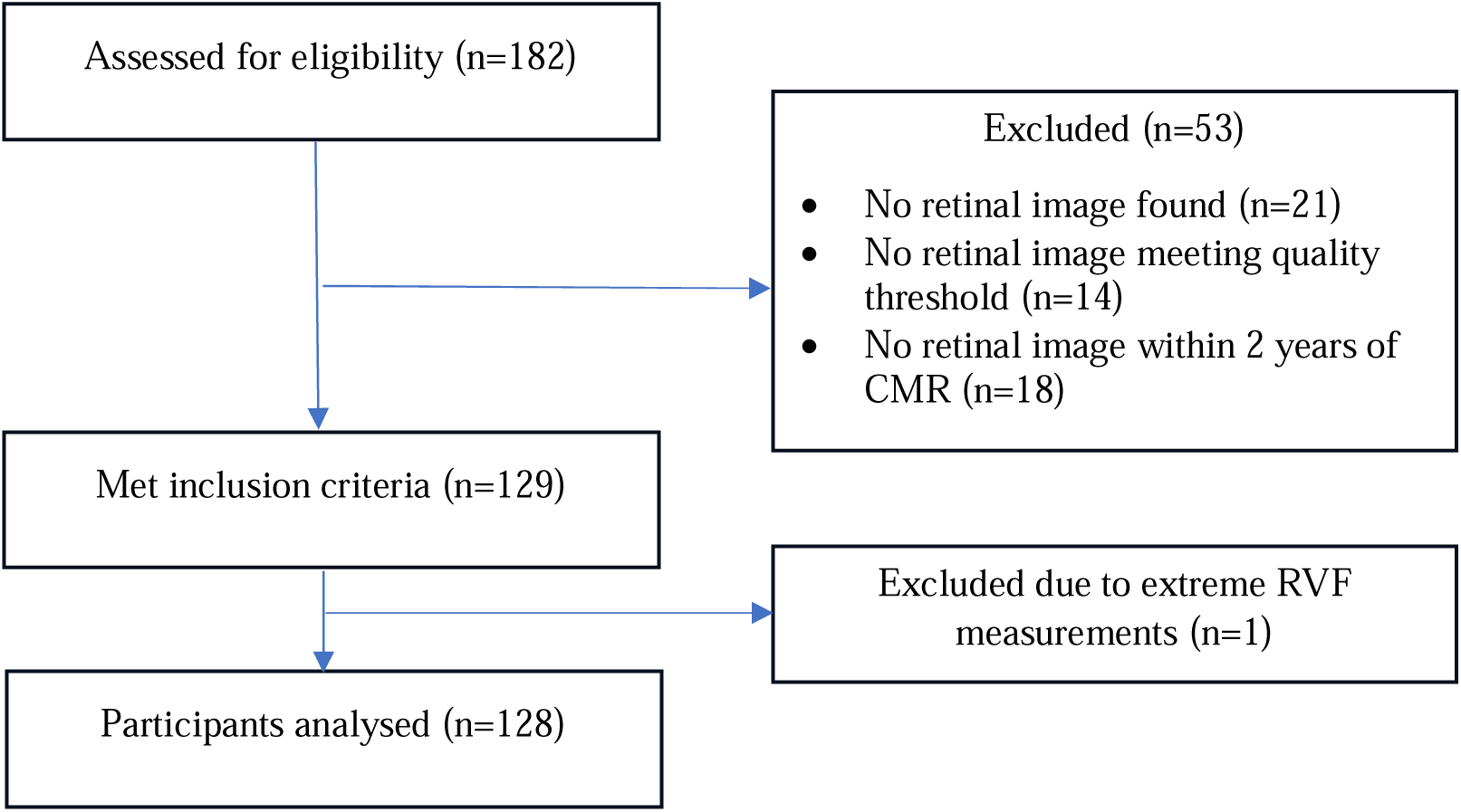
Participant flow diagram.

**Table 1:**
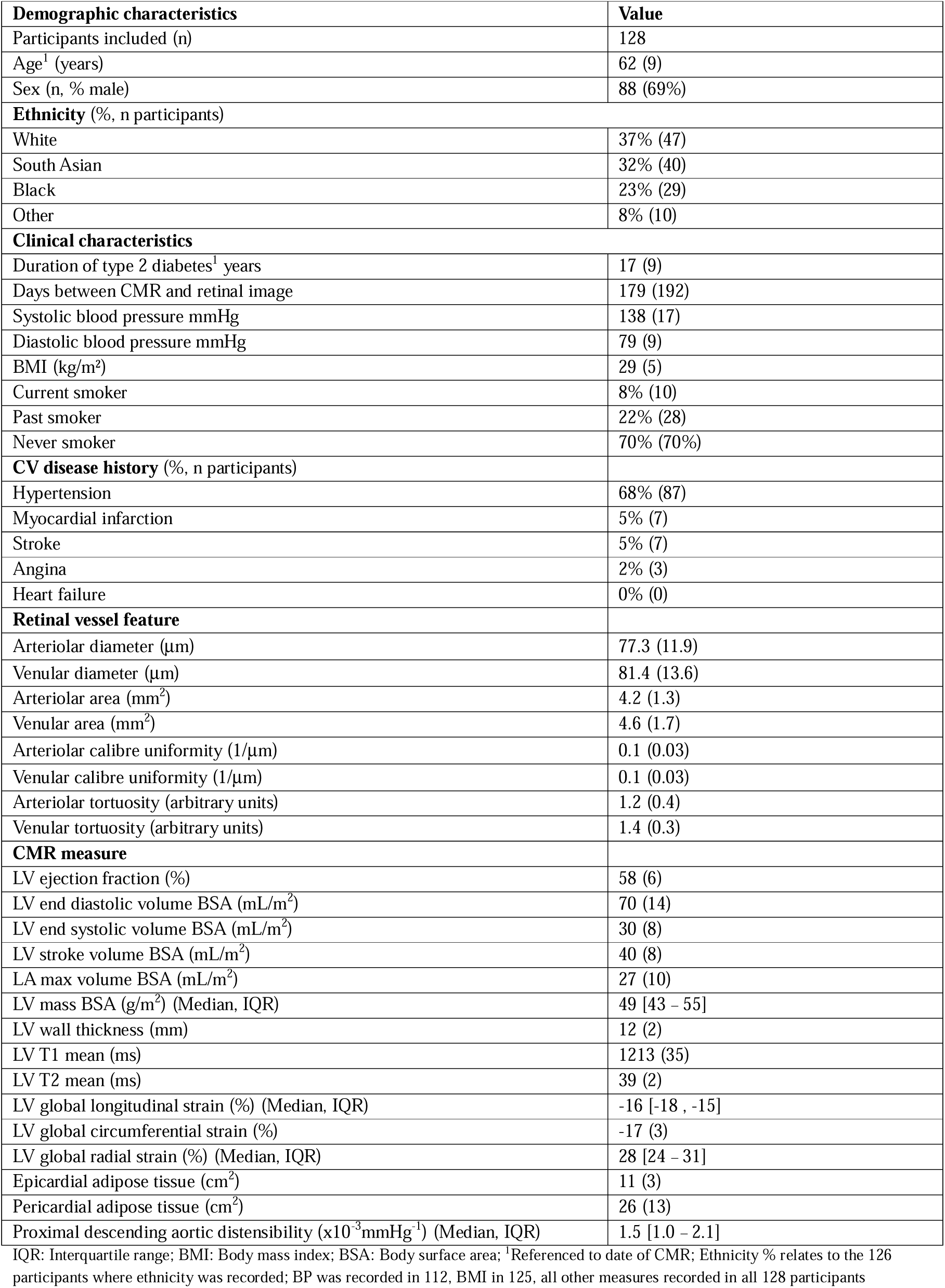
Baseline characteristics of included participants. Mean (SD) unless otherwise stated.

68% of patients had a known diagnosis of hypertension. Previous CV events were rare (5% myocardial infarction, 5% stroke, 2% angina, 0% heart failure), with all events occurring >12 months prior to recruitment.

### CMR and RVF baseline characteristics

Fifteen CMR measures met the threshold of ≥70% reporting completeness and were included for further analysis (table 1; CMR measure reference values are in supplementary table 5).

LV function and volumes were consistent with preserved systolic function (LVEF 58%, LVESVi 30 mL/m^2^, and LVSVi 40mL/m^2^). Normal LVEDVi (70mL/m^2^), LVMi (49g/m^2^), LAVi (27mL/m^2^) and high-normal LVWT (12mm) indicated there was no evidence of adverse chamber remodelling.

Mean LV T1 (1213ms) and T2 (39ms) signals were within a normal range. LV GLS (−16%) and LV GCS (−17%) were mildly impaired, while LV GRS (median 28%) was normal. Mean EAT (11cm^2^) and PAT area (26cm^2^) were normal. Mean proximal descending aortic distensibility (PDAoD) was reduced (1.5 x10^-3^mmHg^-1^), consistent with increased arterial stiffness observed in diabetes^16^.

Participant RVF measures derived from QUARTZ are presented in table 1. Arteriolar diameter (77.3μm) and area (4.2mm^2^) were smaller than venular diameter (81.4μm) and area (4.6mm^2^). Arteriolar calibre uniformity and venular calibre uniformity (0.1 1/μm) were comparable. Tortuosity was lower among arterioles (1.2) than venules (1.4).

### Correlation analysis

Correlation analysis was performed for the 120 included RVF-CMR pairs (supplementary table 6). Overall, 15 of these pairs reached the threshold of |r| ≥0.15 and p<0.05 for inclusion in multivariable regression analysis. Correlations not reaching this threshold included those between RVF measures and LVEF, LVEDVi, LVESVi, LVSVi, LVMi, LVWT, and LV GCS and LV GRS. While the correlation between venular tortuosity and PAT area met threshold for inclusion, the negative directionality was considered to lack biological plausibility and was therefore excluded from subsequent analysis. A further two pairs were selected for regression due to borderline statistical significance and biological plausibility of a potential relationship (venular area and LV GRS, r=0.18, p=0.052; and venular area and LVEF, r=0.15, p=0.097). This resulted in a final selection of 16 RVF-CMR pairs for further analysis with multivariable regression (table 2).

**Table 2:** Correlation & fully adjusted regression models for the 16 RVF-CMR associations.

|  |  | Correlation |  | Fully adjusted regression |  |  |  |
| --- | --- | --- | --- | --- | --- | --- | --- |
| Predictor RVF variable | Outcome CMR variable | r | p-value | $\beta$ | $\beta_{SD}$ | p-value | n |
| Arteriolar diameter | LV T1 mean (ms) | 0.19 | 0.041 | 5.6 (-0.91 - 12.17) | 0.16 | 0.091 | 120 |
| Arteriolar diameter | LV T2 mean (ms) | 0.20 | 0.027 | 0.3 (-0.13 - 0.64) | 0.11 | 0.197 | 123 |
| Arteriolar area | PDAoD ($\times 10^{-3} \text{mmHg}^{-1}$) | 0.28 | 0.008 | 0.2 (-0.01 - 0.36) | 0.18 | 0.057 | 89 |
| Venular area | LV Ejection Fraction | 0.15 | 0.097 | 0.9 (-0.18 - 1.96) | 0.15 | 0.102 | 121 |
| Venular area | LV Global Radial Strain (%) | 0.18 | 0.052 | 1.1 (-0.03 - 2.29) | 0.17 | 0.056 | 114 |
| Venular area | PDAoD ($\times 10^{-3} \text{mmHg}^{-1}$) | 0.25 | 0.017 | 0.2 (0.02 - 0.38)* | 0.19* | 0.032 | 89 |
| Arteriolar CU | LV T1 mean (ms) | -0.25 | 0.006 | -8.8 (-17.35 , -0.21)* | -0.25* | 0.045 | 120 |
| Arteriolar CU | LV T2 mean (ms) | -0.25 | 0.006 | -0.1 (-0.62 - 0.43) | -0.04 | 0.717 | 123 |
| Venular CU | LV T1 mean (ms) | -0.21 | 0.021 | -6.2 (-14.95 - 2.51) | -0.18 | 0.161 | 120 |
| Venular CU | LV T2 mean (ms) | -0.28 | 0.002 | -0.1 (-0.67 - 0.39) | -0.06 | 0.594 | 123 |
| Venular CU | PDAoD ($\times 10^{-3} \text{mmHg}^{-1}$) | 0.24 | 0.027 | 0.1 (-0.15 - 0.33) | 0.09 | 0.462 | 89 |
| Arteriolar tortuosity | Epicardial adipose tissue ($\text{cm}^2$) | 0.19 | 0.046 | 0.3 (-0.33 - 0.97) | 0.10 | 0.331 | 115 |
| Venular tortuosity | Left atrial volume ($\text{ml}/\text{m}^2$) | 0.21 | 0.02 | 1.9 (0.07 - 3.78)* | 0.20* | 0.042 | 121 |
| Venular tortuosity | LV T1 mean (ms) | 0.19 | 0.042 | 5.9 (-0.19 - 12.02) | 0.17 | 0.058 | 120 |
| Venular tortuosity | LV T2 mean (ms) | 0.25 | 0.005 | 0.5 (0.10 - 0.82)* | 0.20* | 0.013 | 123 |
| Venular tortuosity | LV Global Longitudinal Strain (%) | 0.19 | 0.047 | 0.6 (0.19 - 1.08)** | 0.26** | 0.006 | 114 |

CU: Calibre uniformity; PDAoD: proximal descending aortic distensibility; $\beta$ represents change in CMR measure units per 1 standard deviation [SD] increase in RVF in fully adjusted regression model 6; $\beta_{SD}$ represents the standard deviation change in the CMR measure per 1-SD increase in RVF in fully adjusted regression model 6; \*p<0.05 \*\*p<0.01

### Model diagnostics and missing data

Residuals from three RVF-CMR pairs involving PDAoD showed marginal normality violations, with robust standard errors applied for these regressions. The MAR assumption was well supported with no systematic differences between participants with and without recorded SBP (supplementary tables 7-8).

### Multivariable regression

Five of the 16 RVF-CMR pairs revealed statistically significant associations in fully adjusted multivariable linear regression models (table 2). Venular tortuosity was the RVF most consistently associated with CMR measures. Each 1-SD increase in venular tortuosity was associated with a 1.9mL (95% confidence interval, 0.1 - 3.8mL) increase in LAVi, 0.6% (0.2 - 1.1%) worsening of LV GLS, and 0.5ms (0.1 - 0.8ms) increase in LV T2 mean. Each 1-SD increase in venular area was associated with a 0.2 x10^-3^mmHg^-1^ (0.02 - 0.4 x10^-3^mmHg^-1^) increase in PDAoD. Arteriolar calibre uniformity was the only non-venular RVF measure that had a statistically significant association with a CMR measure. Each 1-SD increase in arteriolar calibre uniformity was associated with an 8.8ms (−17.4, - 0.2ms) decrease in LV T1 mean. No significant associations were found in regressions involving LVEF (with venular area, p=0.102) and EAT (with arteriolar tortuosity, p=0.331). Arteriolar diameter, venular calibre uniformity and arteriolar tortuosity were RVF predictors not found to be significantly associated with any of the outcome CMR variables. Changes in SD units of CMR measure per 1-SD increase in RVF (βSD) indicated a moderate magnitude of effect for the five significant associations (figure 2).

**Figure 2:**
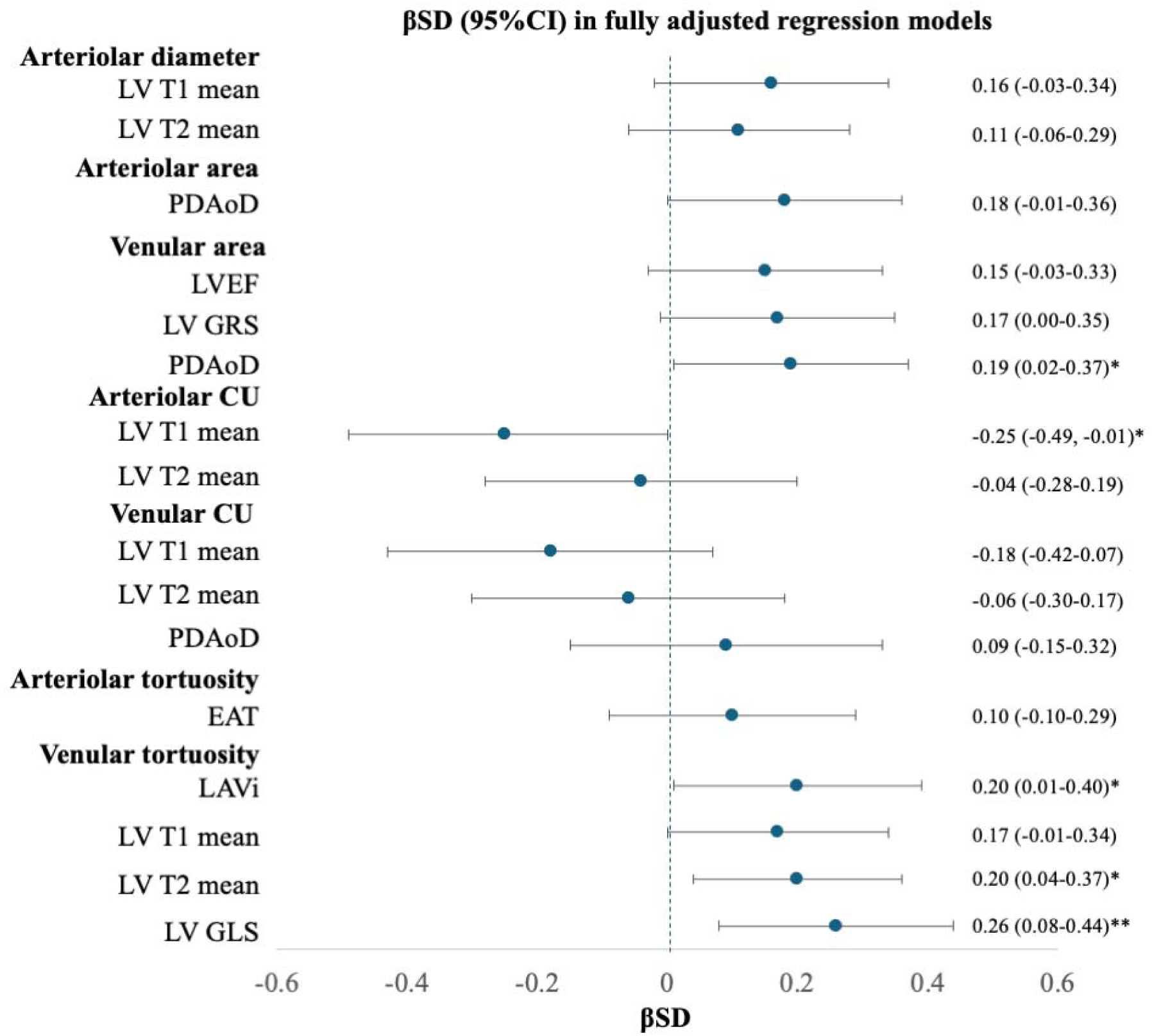
Standardised regression coefficient (βSD) for the 16 RVF-CMR associations in fully adjusted models. PDAoD: proximal descending aortic distensibility; LVEF: Left ventricle ejection fraction; LV GRS: Left ventricle global radial strain; EAT: Epicardial adipose tissue; LAVi: Left atrial max volume indexed to BSA; LV GLS: Left ventricle global longitudinal strain; Forest plot demonstrates standardised regression coefficients (βSD) with 95% confidence intervals for the association between each RVF and CMR measure in the fully adjusted model 6. βSD represents the standard deviation change in the CMR measure per 1-SD increase in RVF in the fully adjusted model 6. Dashed vertical line represents the null (βSD=0); *p<0.05 **p<0.01.

Relatively modest median changes in the β regression coefficient estimates and no change in directionality of associations were observed across sequential models (supplementary table 9). Sensitivity analysis assessing coefficient stability under reduced sample size showed generally low coefficients of variation (median 19%), suggesting the primary significant associations identified through multivariable regression were robust (supplementary table 10).

## Discussion

In this cohort of patients with type 2 diabetes, we demonstrate a range of novel and biologically coherent associations between measures of the retinal microvasculature and subclinical features of CV dysfunction. To our knowledge, this is the first study to identify several significant associations between AI-derived measures of the retinal microvasculature and comprehensive CMR phenotypes.

Venular tortuosity was significantly associated with LV T2 mean, LV global longitudinal strain and left atrial volume. These CMR measures are consistent with a pattern of early diffuse myocardial dysfunction from low grade myocardial inflammation (increased T2), to subclinical impairment of LV contractility (less negative values of longitudinal strain) and atrial remodelling (increased LA volume)^17,18,19^. The biological coherence of our findings is strengthened by the fact that this constellation of subclinical abnormalities is recognized to precede the development of overt heart failure syndromes^20^. Beyond venular tortuosity, we also noted that greater venular area was associated with increased proximal descending aortic distensibility and had a borderline association with better LV contractility (more positive values of radial strain). The directionality of these relationships aligns with previous findings that smaller retinal venular area is associated with an adverse cardiometabolic profile^21^.

Overall, these associations support hypotheses that retinal venular abnormalities reflect microvascular pathology relevant to early CV disease^22^. While CV risk has traditionally focused on the arterial system, our findings suggest that the retinal venular bed also carries important information regarding CV dysfunction.

We also demonstrated associations between features of the retinal arteriolar bed and CMR measures. Greater arteriolar calibre uniformity was associated with lower LV T1 mean. This is also biologically coherent and suggests that deranged retinal arteriolar morphology may reflect broader systemic fibrotic processes, including myocardial fibrosis. We also observed a borderline association between greater arteriolar area and increases in proximal descending aortic distensibility. This aligns with recent work demonstrating a positive correlation between retinal arteriolar density and aortic distensibility^23^, and reflects known shared pathophysiological links between the microvasculature and large arteries^24^. Together, these findings suggest that both the retinal arteriolar and venular beds can contribute to our understanding of distinct elements of subclinical CV dysfunction.

We did not observe any significant relationships between LV functional or volumetric parameters and RVF measures. This may support the hypothesis that changes in RVF are primarily reflective of early microvascular dysfunction that precedes downstream structural remodelling and functional disease.

In a comparable analysis of AI-derived retinal microvascular geometric features and CMR measures of subclinical CV dysfunction, Alatrany *et al* reported a single association between central retinal venular equivalents and LV mass/volume ratio^8^. In the present study, we identified a broader set of associations, including a coherent venular tortuosity signature encompassing LV T2, LV GLS, and LAVi. An explanation for this may be the distinct methodological approaches taken to quantify geometric features of the retinal microvasculature. Their study derived zone-based calibre equivalents from the largest retinal vessels in the standard zone, whereas our study extracted a richer phenotype through computing vessel-segment level measurements weighted by segment length across the full microvascular tree. Additionally, our study incorporated a QUARTZ derived measure of calibre uniformity, which was a significant predictor in our analysis. Our study also examined a wider panel of CMR measures, including some that we found to be significant outcome variables such as LV T1 and T2 mean, LAVi, and PDAoD. Finally, their end-to-end deep learning models were applied to binary classification of the presence or absence of a set pathological CMR threshold from retinal images. This is a fundamentally different analytic approach to our explicit quantification of RVFs and regression against continuous CMR measures, which may have resulted in a greater statistical power for us to detect significant relationships.

A key strength of this study is that we demonstrated coefficient estimates that are biologically coherent, directionally consistent, and stable across sensitivity analyses, with multivariable regression models accounting for several potential confounders. Another strength was the use of QUARTZ itself, which uses AI to extract a rich retinal vascular phenotype from across whole images of varying quality and has been extensively validated for use in large clinical and population-based studies^14^. The reproducibility and granularity of RVF estimates derived from QUARTZ enabled more extensive and robust exploration of relationships with CMR measures than those reported in other studies^8^. Additionally the type 2 diabetes cohort, while disease-specific, provided sufficient variance in subclinical CMR measures to detect associations while avoiding the potential confounding of established CV disease.

Limitations of this study include its small sample size, which meant we had limited power to detect several potentially true associations. Sensitivity analysis with a ∼10% reduction in sample size indicated that our five statistically significant associations had stable effect sizes and directionality, but significance was sometimes inconsistent across iterations. This highlights the need for a larger study to increase precision of effect size estimates, evaluate robustness of these findings, and determine the potential predictive utility of RVF measures. While this diabetic cohort was appropriate for identifying subclinical cardiac dysfunction, further study should include healthy controls and those with established CV disease. This will test the generalisability of these findings and determine whether the observed associations are diabetes-specific or exist across the spectrum of CV health. Additionally, it is known that retinal vascular phenotype can vary between ethnic groups^25^, and while we recruited a study cohort who were ethnically diverse, our sample size precluded meaningful stratified analysis by ethnicity. Finally, this was a cross-sectional study where the CMR could occur before or after retinal image acquisition. Further prospective studies will enable ascertainment of the temporality of the relationship between RVF and CMR measures and facilitate predictive inferences.

The five significant associations we observed demonstrated a moderate magnitude of effect, with a mean 0.21 SD change in CMR measure per 1-SD increase in RVF measure, consistent with other epidemiological studies examining CMR against demographic and clinical predictors^26^. The moderate absolute effect sizes likely reflect range restriction in a cohort without overt CV disease. In a cohort with established CV disease, where CMR measures would span a greater pathological range, larger and more clinically meaningful effect sizes would be anticipated. However, this may itself inform the most appropriate use cases for retinal vascular imaging. It is less likely this technique would be deployed to finalise the binary presence or absence of CV disease. Rather, ours and others’ findings point more toward use in population health screening where the degree of subclinical CV dysfunction is evaluated to inform prioritisation of further diagnostic investigations, such as CMR or coronary angiography. Beyond atherosclerotic CV disease, our findings suggest potential applications for conditions characterised by microvascular dysfunction such as heart failure with preserved ejection fraction (HFpEF)^27^ and ischaemia with non-obstructed coronary arteries (INOCA)^28^, as well as those at risk of aortic events or inherited cardiomyopathies.

These findings demonstrate that features of the retinal microvasculature are associated with subclinical measures of CV dysfunction detected by CMR in patients with type 2 diabetes. These are novel and biologically coherent associations that build on prior work through use of a validated AI-enabled retinal image analysis tool. Our study establishes a foundation for larger-scale replication and further investigation of these relationships with more directed study design in other patient cohorts. This will enable testing the predictive potential of RVF measures at different stages of CV disease processes and provide new mechanistic insights into the shared pathophysiology of micro- and macrovascular disease. Collectively, our findings strengthen the case for retinal imaging as a scalable and accessible method for detecting and evaluating subclinical CV dysfunction.

## Sources of funding

This study received no specific grant from any funding agency in the public, commercial or not-for-profit sectors. The UKIDS trial is sponsored by Perspectum Ltd. The sponsor was involved in data collection, analysis, and interpretation. Use of the North East London Diabetic Eye Screening programme data was funded by Wellcome Collaborative award (224390/Z/21/Z). CW’s time was funded by the National Institute for Health and Care Research.

## Data availability statement

Anonymised individual patient data can be shared upon request or as required by law and/or regulation with qualified external researchers. Approval of such requests is at the discretion of the study sponsors and is dependent on the nature of the request, the availability of the data, and the intended use of the data.

## Disclosures

MM, HTB, CD, and RP are employees at Perspectum Ltd., the company that developed CoverScan™ used in this study, and funded recruitment of participants to the UKIDS trial. HTB is also a shareholder at Perspectum Ltd. ERB receives grant funding from The British Heart Foundation for projects unrelated to this study.

## Author contributions

CW, ARR, IY, MM, and CGO conceptualised the study. Study methodology was developed by CW, ARR, RAW, JF, SAB, CGO. Investigation was conducted by HED, CZ, IY, RDH, MM, HTB, CD, RP, JA, and RC. Formal analysis and interpretation were performed by CW and MM. The original manuscript was prepared by CW. All authors contributed to editing of the manuscript and approved the final version for submission. Supervision was provided by ARR, MM, ERB, and CGO. CW had full access to all data in the study and takes responsibility for their integrity and the accuracy of the data analysis.

## Notes

### Author Declarations

West Midlands - Solihull research ethics committee gave ethical approval for the UKIDS study (ethics reference 20/WM/0007) NHS Health Research Authority and Health and Care Research Wales gave ethical approval for retinal imaging data used from the North East London Diabetic Eye Screening programme (Integrated Research Application System number 265637)

### Summary of Updates

Updated content with some reframing of the focus of the discussion. No change in the analytic methodology or results.

