## Supplementary appendix for "Retinal microvascular features are associated with CMR measures of subclinical cardiovascular dysfunction"

**Supplementary table 1:** Cardiovascular magnetic resonance (CMR) features reported and completion rate

| **CMR measure** | **Unit of measure** | **Reporting completion (%)** |
|---|---|---|
| Left ventricular ejection fraction (LVEF) | % | 95% |
| Left ventricular end diastolic volume | mL | 95% |
| Left ventricular end diastolic volume BSA (LVEDVi) | $mL/m^2$ | 95% |
| Left ventricular end systolic volume | mL | 95% |
| Left ventricular end systolic volume BSA (LVESVi) | $mL/m^2$ | 95% |
| LV stroke volume | mL | 95% |
| LV stroke volume BSA (LVSVi) | $mL/m^2$ | 95% |
| LA max volume | mL | 95% |
| LA max volume BSA (LAVi) | $mL/m^2$ | 95% |
| LV mass | g | 95% |
| LV mass BSA (LVMi) | $g/m^2$ | 95% |
| LV wall thickness (LVWT) | mm | 95% |
| LV T1 Mean | ms | 94% |
| LV T2 Mean | ms | 96% |
| Epicardial adipose tissue (EAT) | $cm^2$ | 90% |
| Pericardial adipose tissue (PAT) | $cm^2$ | 90% |
| LV global longitudinal strain (LV GLS) | % | 89% |
| LV global circumferential strain (LV GCS) | % | 89% |
| LV global radial strain (LV GRS) | % | 89% |
| Ascending aortic distensibility | $x10^{-3}mmHg^{-1}$ | 49% |
| Abdominal aortic distensibility | $x10^{-3}mmHg^{-1}$ | 59% |
| Proximal descending aortic distensibility (PDAoD) | $x10^{-3}mmHg^{-1}$ | 70% |
| Aortic wall thickness | mm | 51% |

Completion rate is as a percentage of the 128 final included participants who also had an eligible retinal image identified

BSA: Body surface area

LV T1 and T2 mean are the relaxation times averaged across all 16 American Heart Association (AHA) myocardial segments derived from the three short-axis views excluding the apex

**Supplementary table 2:** Demographic and clinical data collected on participants

| **Demographic data** | **Response options** |
|---|---|
| Date of birth | DD-MM-YYYY |
| Sex | Male/Female |
| Ethnicity | White/South Asian/Black/Other |
| Height | cm |
| Weight | kg |
| Waist circumference | cm |
| Hip circumference | cm |
| Smoking status | Current/Past/Never |
| **Clinical data** | |
| Type 2 diabetes | Yes/No |
| Type 2 treatment | Medication/Lifestyle advice/Medication & Lifestyle/Other/None |
| Type 2 diabetes year of diagnosis | XXXX |
| Diabetic retinopathy | Yes/No |
| Hypertension diagnosis | Yes/No/Don't Know |
| Hypertension treatment | Medication/Lifestyle advice/Medication & Lifestyle/Other/None |
| High cholesterol diagnosis | Yes/No/Don't Know |
| High cholesterol treatment | Medication/Lifestyle advice/Medication & Lifestyle/Other/None |
| Myocardial infarction | Yes/No |
| Angina | Yes/No |
| Atrial fibrillation/other arrhythmia | Yes/No |
| Heart failure | Yes/No |
| Coronary artery bypass grafting | Yes/No |
| Percutaneous coronary intervention | Yes/No |
| Cerebrovascular event | Yes/No |
| Systolic blood pressure | $mmHg^{-3}$ |
| Diastolic blood pressure | $mmHg^{-3}$ |

**Supplementary table 3:** Additional details on statistical methods

| Statistical method | Further details |
|---|---|
| a. Normality assessment | For each RVF-CMR pair of interest, we interrogated normality assumptions through examining the residual distributions of the outcome variables. We performed visual inspection of residual histograms and Q-Q plots, performed Shapiro-Wilk test ($p<0.05$), examined skewness (threshold of >2), and kurtosis (threshold of <0 or >6). Where evidence of non-normality was found, the CMR outcome was log transformed and re-tested to see whether this improved normality of distribution. Where normality was not improved, robust standard error was used for regression. Output of this analysis is in supplementary table 7. |
| b. Missing at Random (MAR) assessment for systolic blood pressure missingness | Compared 112 participants with SBP vs. 16 participants without SBP recorded for significant differences between key characteristics using t-tests for continuous variables and chi-square tests for categorical variables ($\alpha=0.05$). The characteristics compared were demographic (age at CMR, duration of diabetes, time between retinal image and CMR, sex, imaging centre), retinal vascular features (arteriolar and venular calibre uniformity, width, and tortuosity), and CMR measures (LV T1 mean, LV T2 mean, LA max volume BSA, LV global longitudinal, radial, and circumferential strain). Output of this analysis is in supplementary table 8. |
| c. Multiple imputation of missing systolic blood pressure (SBP) data | Twenty imputed datasets (m=20) were generated using k=5 nearest neighbours with random seeding. The imputation model used variables with known associations with SBP, which included age, sex, duration of diabetes, and time interval between retinal and CMR imaging. The algorithm identified 5 participants with observed SBP most similar to each participant with unrecorded SBP to generate 20 plausible SBP values per participant. Rubin's rules were then applied to pool coefficients across each regression model fitted to these 20 datasets to obtain final SBP estimates for participants where SBP had been unrecorded. |
| d. Construction of imaging centre variable ($\alpha_j$) | Due to highly unbalanced sample sizes across imaging centres (range 1-71), centres were collapsed into a binary variable of centre 1 (n=71) vs. other (n=57), the latter of which was composed of participants from centres 2-5. |
| e. Sensitivity analysis to reduced sample | We randomly removed n=13 (representing ~10% of the final participant sample size) from each of the final RVF-CMR multivariable regression pairings and reran the regressions through 100 iterations. Mean $\beta$ and $\beta$std were calculated across each of the 100 iterations for each regression pair. Coefficient of variation was calculated as well as the proportion of iterations where the coefficient estimate was found to be statistically significant ($\alpha=0.05$), termed the rate of significance. Output of this analysis is in supplementary table 10. |

**Supplementary table 4:** Regression models specification

| Model number | Regression equation |
|---|---|
| 1 | $Y = \beta_0 + \beta_1(RVF) + \varepsilon$ |
| 2 | $Y = \beta_0 + \beta_1(RVF) + \beta_2(Age) + \varepsilon$ |
| 3 | $Y = \beta_0 + \beta_1(RVF) + \beta_2(Age) + \beta_3(Sex) + \varepsilon$ |
| 4 | $Y = \beta_0 + \beta_1(RVF) + \beta_2(Age) + \beta_3(Sex) + \beta_4(SBP) + \varepsilon$ |
| 5 | $Y = \beta_0 + \beta_1(RVF) + \beta_2(Age) + \beta_3(Sex) + \beta_4(SBP) + \beta_5(T2Duration) + \beta_6(Days) + \varepsilon$ |
| 6 | $Y = \beta_0 + \beta_1(RVF) + \beta_2(Age) + \beta_3(Sex) + \beta_4(SBP) + \beta_5(T2Duration) + \beta_6(Days) + \alpha_j + \varepsilon$ |

Y: CMR measure; RVF: Retinal vessel feature; Age: age at CMR (years); Sex: 1 Male, 2 Female; SBP: Systolic blood pressure; T2Duration: Duration of type 2 diabetes from diagnosis to CMR date; Days: Interval between retinal and CMR image acquisition (days); $\varepsilon$= residual error term; $\alpha_j$ = imaging centre fixed effect

**Supplementary table 5**: CMR measure reference values

| CMR measure | Reference range | Source |
|---|---|---|
| LVEF | ≥50% | McDonagh TA et al (2023)[1] |
| LVEDVi* | Male: 60 - 109 ml/m$^2$<br>Female: 56 - 96 ml/m$^2$<br>Asian Male: 53 – 92 ml/m$^2$<br>Asian Female: 50 – 90 ml/m$^2$ | Zhan Y et al (2024)[2] |
| LVESVi* | Male: 18 – 45 ml/m$^2$<br>Female: 16 – 38 ml/m$^2$<br>Asian Male: 15 - 38 ml/m$^2$<br>Asian Female: 14 – 33 ml/m$^2$ | Zhan Y et al (2024)[2] |
| LVSVi* | Male: 36 – 69 ml/m$^2$<br>Female: 35 – 60 ml/m$^2$<br>Asian Male: 35 – 62 ml/m$^2$<br>Asian Female: 35 – 61 ml/m$^2$ | Zhan Y et al (2024)[2] |
| LAVi | Male: < 59 ml/m$^2$<br>Female: < 60 ml/m$^2$ | Kawel-Boehm N et al (2020)[3] |
| LVMi* | Male: < 76 g/m$^2$<br>Female: < 57 g/m$^2$<br>Asian Male: < 71 g/m$^2$<br>Asian Female: < 56 g/m$^2$ | Zhan Y et al (2024)[2] |
| LVWT | Male: ≤ 12mm<br>Female: ≤ 10 mm | Kawel-Boehm N et al (2020)[3] |
| LV T1 mean | 3T: < 1269ms | Roca-Fernandez A et al (2023)[4] |
| LV T2 mean | 3T: < 46ms | Roca-Fernandez A et al (2023)[4] |
| LV GLS | Male: < -14.8%<br>Female: < -16.4% | 97.5$^{th}$ percentile from COVERSCAN study[5] |
| LV GCS | Male: < -15%<br>Female: < -15.1% | 97.5$^{th}$ percentile from COVERSCAN study[5] |
| LV GRS | Male: <36.2%<br>Female: <42.5% | 97.5$^{th}$ percentile from COVERSCAN study[5] |
| EAT | Male: < 15.1 cm$^2$<br>Female: <12.2 cm$^2$ | Mean ±1.96SD[5] |
| PAT | Male: < 37.6 cm$^2$<br>Female: < 29.5 cm$^2$ | Male: Mean ±1.96SD[5]<br>Female: 97.5$^{th}$ percentile[5] |
| PDAoD | Male: >2.91 x10$^{-3}$mmHg$^{-1}$<br>Female: >2.11 x10$^{-3}$mmHg$^{-1}$ | 97.5$^{th}$ percentile from COVERSCAN study[5] |

*Asian refers to South Asian (Indian, Pakistani, Bangladeshi, or other South Asian) or Chinese ethnicity

**Supplementary Table 6:** Pearson correlation matrix for RVF vs. CMR variables

| CMR variable (n) | Arteriolar diameter | Venular diameter | Arteriolar area | Venular area | Arteriolar calibre uniformity | Venular calibre uniformity | Arteriolar tortuosity | Venular tortuosity |
|---|---|---|---|---|---|---|---|---|
| LVEF (121) | 0.03 | 0.05 | 0.09 | 0.15 | 0.13 | 0.03 | -0.16 | -0.06 |
| LVEDVi (121) | 0.09 | 0.06 | 0.01 | -0.09 | -0.00 | 0.04 | 0.05 | 0.01 |
| LVESVi (121) | 0.05 | 0.02 | -0.04 | -0.13 | -0.07 | 0.02 | 0.10 | 0.04 |
| LVSVi (121) | 0.10 | 0.09 | 0.05 | -0.02 | 0.06 | 0.06 | -0.02 | -0.02 |
| LAVi (121) | 0.11 | 0.06 | -0.06 | -0.09 | 0.03 | -0.01 | 0.03 | **0.21*** |
| LVMi (121) | 0.03 | 0.03 | -0.01 | -0.10 | -0.06 | -0.04 | 0.11 | 0.08 |
| LVWT (121) | 0.00 | -0.02 | -0.05 | -0.11 | -0.06 | -0.05 | 0.11 | 0.07 |
| LV T1 mean (120) | **0.19*** | 0.17 | -0.12 | -0.16 | **-0.25**** | **-0.21*** | 0.15 | **0.19*** |
| LV T2 mean (123) | **0.20*** | 0.18 | -0.03 | -0.01 | **-0.25**** | **-0.28**** | 0.07 | **0.25**** |
| LV GLS (114) | -0.10 | -0.16 | -0.17 | -0.12 | -0.03 | 0.05 | 0.03 | **0.19*** |
| LV GCS (114) | -0.02 | -0.08 | -0.06 | -0.16 | -0.13 | -0.03 | 0.15 | 0.05 |
| LV GRS (114) | 0.02 | 0.08 | 0.09 | 0.18 | 0.13 | 0.05 | -0.15 | -0.02 |
| EAT (115) | 0.04 | 0.01 | -0.08 | -0.12 | -0.13 | -0.16 | **0.19*** | -0.03 |
| PAT (115) | 0.07 | 0.12 | -0.04 | 0.02 | -0.17 | -0.16 | 0.15 | **-0.20*** |
| PDAoD (89) | 0.07 | 0.04 | **0.28**** | **0.25*** | 0.21 | **0.24*** | -0.01 | -0.01 |

Values refer to Pearson's correlation coefficient
i denotes metric indexed to body surface area
n denotes the # of participants in whom the CMR variable was recorded and therefore were included in the pairwise correlation
*p<0.05 **p<0.01; bold denotes significant correlations at α=0.05

**Supplementary Table 7:** Normality assessment for variables in adjusted regression models

| Predictor RVF variable | Outcome CMR variable | Sample | Shapiro-Wilk | p-value | Skewness | Kurtosis | Outcome | Log transform outcome |
|---|---|---|---|---|---|---|---|---|
| Arteriolar diameter (mm) | LV T1 mean (ms) | 105 | 0.99 | 0.384 | 0.04 | 2.46 | Normal | N/A |
| Arteriolar diameter (mm) | LV T2 mean (ms) | 108 | 0.99 | 0.856 | 0.08 | 3.27 | Normal | N/A |
| Arteriolar area (mm²) | PDAoD ($x10^{-3}mmHg^{-1}$) | 79 | 0.97 | 0.040 | 0.64 | 4.83 | Non-normal | No log improvement |
| Venular area (mm²) | LV ejection fraction (%) | 105 | 0.99 | 0.359 | 0.34 | 2.88 | Normal | N/A |
| Venular area (mm²) | LV GRS (%) | 98 | 0.99 | 0.671 | 0.07 | 3.36 | Normal | N/A |
| Venular area (mm²) | PDAoD ($x10^{-3}mmHg^{-1}$) | 79 | 0.96 | 0.025 | 0.66 | 5.06 | Non-normal | No log improvement |
| Arteriolar calibre uniformity (1/mm) | LV T1 mean (ms) | 105 | 0.99 | 0.525 | 0.09 | 2.52 | Normal | N/A |
| Arteriolar calibre uniformity (1/mm) | LV T2 mean (ms) | 108 | 0.99 | 0.621 | 0.12 | 3.36 | Normal | N/A |
| Venular calibre uniformity (1/mm) | LV T1 mean (ms) | 105 | 0.99 | 0.490 | 0.07 | 2.52 | Normal | N/A |
| Venular calibre uniformity (1/mm) | LV T2 mean (ms) | 108 | 0.99 | 0.680 | 0.10 | 3.36 | Normal | N/A |
| Venular calibre uniformity (1/mm) | PDAoD ($10^{-3}mmHg^{-1}$) | 79 | 0.97 | 0.033 | 0.60 | 4.45 | Non-normal | No log improvement |
| Arteriolar tortuosity | EAT (cm²) | 99 | 0.97 | 0.051 | 0.61 | 3.58 | Normal | N/A |
| Venular tortuosity | LAVi (ml/m²) | 106 | 0.98 | 0.174 | 0.45 | 2.92 | Normal | N/A |
| Venular tortuosity | LV T1 mean (ms) | 105 | 0.99 | 0.416 | -0.17 | 2.59 | Normal | N/A |
| Venular tortuosity | LV T2 mean (ms) | 108 | 0.99 | 0.664 | 0.10 | 3.35 | Normal | N/A |
| Venular tortuosity | LV GLS (%) | 98 | 0.99 | 0.867 | -0.03 | 2.77 | Normal | N/A |

**Supplementary Table 8:** Missing at random assessment (MAR) for participants where SBP is missing vs. SBP imputed 16 sample

| | | Participants with BP readings | | | Participants without BP readings | | | |
|---|---|---|---|---|---|---|---|---|
| | | Value | n | SD/IQR | Value | n | SD/IQR | P value |
| Demographics/Clinical | Sex (Female/Male) | 34/78 | 112 | n/a | 6/10 | 16 | n/a | 0.56 |
| | BMI (kg/m²) | 29.4 | 112 | 4.8 | 29.8 | 12 | 5.8 | 0.76 |
| | Days between images | 190.5 | 112 | 198.2 | 94.8 | 16 | 118.2 | 0.06 |
| | Diabetes duration (years) | 17.4 | 112 | 9.0 | 14.8 | 16 | 10.0 | 0.30 |
| | Age at CMRI (years) | 62.3 | 112 | 9.2 | 63.2 | 16 | 11.0 | 0.71 |
| Retinal Vascular Feature | Arteriolar diameter (μm) | 76.8 | 112 | 11.1 | 80.4 | 16 | 16.6 | 0.27 |
| | Venular diameter (μm) | 80.7 | 112 | 12.9 | 86.2 | 16 | 17.4 | 0.13 |
| | Arteriolar area (mm²) | 4.2 | 112 | 1.4 | 4.3 | 16 | 0.9 | 0.73 |
| | Venular area (mm²) | 4.5 | 112 | 1.8 | 5.1 | 16 | 1.4 | 0.17 |
| | Arteriolar calibre uniformity (1/μm) | 0.1 | 112 | 0.03 | 0.10 | 16 | 0.03 | 0.32 |
| | Venular calibre uniformity (1/μm) | 0.1 | 112 | 0.03 | 0.11 | 16 | 0.03 | 0.18 |
| | Arteriolar tortuosity | 1.2 | 112 | 0.4 | 1.30 | 16 | 0.5 | 0.33 |
| | Venular tortuosity | 1.4 | 112 | 0.3 | 1.52 | 16 | 0.4 | 0.05 |
| CMR features | LVEF (%) | 57.5 | 105 | 5.9 | 58.8 | 16 | 5.8 | 0.45 |
| | LAVi (ml/m²) | 25.3 | 106 | 19.8 – 33.2 | 28.6 | 15 | 22.1 – 34.6 | 0.23 |
| | LV Mass BSA (g/m²) | 49.4 | 105 | 43.4 – 55.4 | 49.2 | 16 | 42.6 – 54.5 | 0.79 |
| | LV T1 Mean (ms) | 1212.5 | 105 | 34.0 | 1219.0 | 15 | 45.4 | 0.51 |
| | LV T2 Mean (ms) | 39.0 | 108 | 2.1 | 40.0 | 15 | 2.7 | 0.08 |
| | EAT | 10.4 | 99 | 3.3 | 10.90 | 16 | 3.66 | 0.60 |
| | PAT | 25.6 | 99 | 12.5 | 27.03 | 16 | 12.72 | 0.68 |
|  | LV GLS (%) | -16 | 98 | -18 , -15 | -16 | 16 | -17 -14.5 | 0.14 |
| | LV GCS (%) | -17.2 | 98 | 2.50 | -17.88 | 16 | 2.47 | 0.34 |
| | LV GRS (%) | 28 | 98 | 24 – 31 | 29 | 16 | 25 – 35 | 0.29 |
| | PDAoD | 1.5 | 79 | 1.0 – 2.1 | 1.7 | 10 | 1.0 – 2.3 | 0.65 |

Median [IQR] comparison with a Mann-Whitney U test for LAVi, LV GLS, LV GRS, and PDAoD
Median [SD] comparison with independent samples t-test for all other retinal vascular feature and CMR feature variables

**Supplementary Table 9:** Full regression model outputs

| | | Model 1 | | Model 2 | | Model 3 | | Model 4 | | Model 5 | | Model 6 | | | |
|---|---|---|---|---|---|---|---|---|---|---|---|---|---|---|---|
| **Predictor RVF variable** | **Outcome CMR variable** | **β (CI)** | **$\beta_{SD}$** | **β (CI)** | **$\beta_{SD}$** | **β (CI)** | **$\beta_{SD}$** | **β (CI)** | **$\beta_{SD}$** | **β (CI)** | **$\beta_{SD}$** | **Β (CI)** | **$\beta_{SD}$** | **p6** | **n** |
| **Arteriolar diameter** | **LV T1 mean (ms)** | 6.5 (0.28 - 12.78) | **0.18*** | 5.9 (-0.36 - 12.11) | 0.17 | 5.9 (-0.39 - 12.13) | 0.17 | 5.9 (-0.13 - 11.97) | 0.17 | 5.6 (-0.47 - 11.59) | 0.16 | 5.6 (-0.91 - 12.17) | 0.16 | 0.091 | 120 |
| **Arteriolar diameter** | **LV T2 mean (ms)** | 0.4 (0.05 - 0.83) | **0.20*** | 0.3 (-0.03 - 0.69) | 0.15 | 0.3 (-0.03 - 0.69) | 0.15 | 0.3 (-0.03 - 0.69) | 0.15 | 0.3 (-0.03 - 0.69) | 0.15 | 0.3 (-0.13 - 0.64) | 0.11 | 0.197 | 123 |
| **Arteriolar area** | **PDAoD ($x10^{-3}mmHg^{-1}$)** | 0.3 (0.08 - 0.49) | **0.28**** | 0.2 (0.00 - 0.34) | 0.17 | 0.2 (0.01 - 0.34) | 0.16 | 0.2 (0.02 - 0.34) | 0.15 | 0.2 (0.02 - 0.34) | 0.16 | 0.2 (0.01 - 0.36) | 0.18 | 0.057 | 89 |
| **Venular area** | **LVEF** | 0.9 (-0.16 - 1.94) | 0.15 | 0.9 (-0.15 - 1.95) | 0.15 | 1.0 (-0.05 - 2.03) | 0.17 | 0.9 (-0.2 - 1.9) | 0.15 | 0.9 (-0.12 – 2.00) | 0.16 | 0.9 (-0.18 - 1.96) | 0.15 | 0.102 | 121 |
| **Venular area** | **LV GRS (%)** | 1.18 (-0.01 - 2.38) | 0.18 | 1.2 (0.06 - 2.41) | **0.19*** | 1.3 (0.17 - 2.45) | **0.20*** | 1.2 (0.07 - 2.38) | **0.19*** | 1.3 (0.11 - 2.46) | **0.20*** | 1.1 (-0.03 - 2.29) | 0.17 | 0.056 | 114 |
| **Venular area** | **PDAoD ($x10^{-3}mmHg^{-1}$)** | 0.3 (0.05 - 0.47) | **0.26*** | 0.2 (0.03 - 0.37) | **0.20*** | 0.2 (0.03 - 0.37) | **0.20*** | 0.2 (0.02 - 0.37) | **0.19*** | 0.2 (0.02 - 0.37) | **0.19*** | 0.2 (0.02 - 0.38)* | **0.19*** | 0.032 | 89 |
| **Arteriolar calibre uniformity** | **LV T1 mean (ms)** | -8.7 (-14.83 , -2.58) | **-0.25**** | -7.5 (-13.92 , -0.99) | **-0.21*** | -7.5 (-13.95 , -0.97) | **-0.21*** | -6.6 (-12.83 , -0.43) | **-0.19*** | -6.0 (-12.25 - 0.22) | -0.17 | -8.8 (-17.35 , -0.21)* | **-0.25*** | 0.045 | 120 |
| **Arteriolar calibre uniformity** | **LV T2 mean (ms)** | -0.5 (-0.92 , -0.16) | **-0.24**** | -0.3 (-0.67 -0.11) | -0.12 | -0.3 (-0.66 - 0.10) | -0.13 | -0.3 (-0.65 - 0.11) | -0.12 | -0.3 (-0.65 - 0.12) | -0.12 | -0.1 (-0.62 - 0.43) | -0.04 | 0.717 | 123 |
| **Venular calibre uniformity** | **LV T1 mean (ms)** | -7.4 (-13.63 , -1.13) | **-0.21*** | -5.8 (-12.54 - 0.85) | -0.17 | -5.8 (-12.58 - 0.90) | -0.16 | -5.5 (-11.89 - 0.91) | -0.16 | -4.8 (-11.24- 1.64) | -0.14 | -6.2 (-14.95 - 2.51) | -0.18 | 0.161 | 120 |
| **Venular calibre uniformity** | **LV T2 mean (ms)** | -0.6 (-1.00 , -0.24) | **-0.28**** | -0.3 (-0.72 - 0.06) | -0.15 | -0.31 (-0.70 - 0.08) | -0.14 | -0.3 (-0.69 - 0.08) | -0.14 | -0.3 (-0.69 - 0.09) | -0.14 | -0.1 (-0.67 - 0.39) | -0.06 | 0.594 | 123 |
| **Venular calibre uniformity** | **PDAoD ($x10^{-3}mmHg^{-1}$)** | 0.2 (0.03 - 0.45) | **0.23*** | 0.0 (-0.15 - 0.21) | 0.03 | 0.0 (0.16 - 0.21) | 0.024 | 0.0 (0.17 - 0.20) | 0.01 | 0.0 (-0.17 - 0.20) | 0.014 | 0.1 (-0.15 - 0.33) | 0.088 | 0.462 | 89 |
| **Arteriolar tortuosity** | **EAT ($cm^2$)** | 0.6 (0.01 - 1.26) | **0.19*** | 0.6 (0.02 - 1.27) | **0.19*** | 0.7 (0.03 - 1.27) | **0.19*** | 0.63 (0.01 - 1.25) | **0.19*** | 0.6 (-0.07 - 1.18) | 0.17 | 0.3 (-0.33 - 0.97) | 0.10 | 0.331 | 115 |
| **Venular tortuosity** | **LAVi ($ml/m^2$)** | 2.1 (0.30 - 3.90) | **0.22*** | 2.1 (0.31 - 3.91) | **0.22*** | 2.0 (0.14 - 3.79) | **0.21*** | 1.9 (0.11 - 3.77) | **0.20*** | 1.9 (0.09 - 3.77) | **0.20*** | 1.9 (0.07 - 3.78)* | **0.20*** | 0.042 | 121 |
| **Venular tortuosity** | **LV T1 mean (ms)** | 6.5 (0.25 - 12.73) | **0.18*** | 6.2 (0.02 - 12.4) | **0.18*** | 6.4 (0.05 - 12.76) | **0.18*** | 6.1 (-0.01 - 12.18) | 0.17 | 5.9 (-0.12 - 11.99) | 0.17 | 5.9 (-0.19 - 12.02)† | 0.17 | 0.058 | 120 |
| **Venular tortuosity** | **LV T2 mean (ms)** | 0.6 (0.18 - 0.95) | **0.25**** | 0.5 (0.17 - 0.88) | **0.23**** | 0.5 (0.11 - 0.83) | **0.21*** | 0.5 (0.10 - 0.82) | **0.21*** | 0.5 (0.09 - 0.82) | **0.20*** | 0.5 (0.10 - 0.82)* | **0.20*** | 0.013 | 123 |
| **Venular tortuosity** | **LV GLS (%)** | 0.5 (0.01 - 0.93) | **0.19*** | 0.5 (0.00 – 0.93) | **0.19*** | 0.7 (0.21 - 1.10) | **0.27**** | 0.6 (0.20 - 1.08) | **0.26**** | 0.6 (0.20 - 1.08) | **0.26**** | 0.6 (0.19 - 1.08)** | **0.26**** | 0.006 | 114 |

*p<0.05 **p<0.01; bold denotes statistically significant coefficients at α=0.05
All confidence intervals (CI) are 95%

**Supplementary table 10:** Sensitivity analysis comparing complete sample multivariable regression output versus average output for smaller samples

| Predictor RVF variable | Outcome CMR variable | Complete sample β | p-value | Average β across sensitivities | Standard deviation β across sensitivities | Coefficient of variation of β | % of β coefficients where p<0.05 |
|---|---|---|---|---|---|---|---|
| Arteriolar diameter | LV T1 mean (ms) | 5.63 | 0.091 | 5.63 | 1.07 | 19% | 12% |
| Arteriolar diameter | LV T2 mean (ms) | 0.26 | 0.197 | 0.25 | 0.07 | 29% | 3% |
| Arteriolar area | PDAoD ($10^{-3}$mmHg$^{-1}$) | 0.18 | 0.057 | 0.18 | 0.03 | 14% | 31% |
| Venular area | LVEF | 0.89 | 0.102 | 0.92 | 0.17 | 19% | 12% |
| Venular area | PDAoD ($10^{-3}$mmHg$^{-1}$) | 0.20* | 0.032 | 0.20 | 0.03 | 13% | 62% |
| Venular area | LV GRS (%) | 1.13 | 0.056 | 1.12 | 0.23 | 20% | 29% |
| Arteriolar calibre uniformity | LV T1 mean (ms) | -8.78* | 0.045 | -8.77 | 1.27 | 15% | 42% |
| Arteriolar calibre uniformity | LV T2 mean (ms) | -0.10 | 0.717 | -0.09 | 0.09 | 98% | 0% |
| Venular calibre uniformity | LV T1 mean (ms) | -6.22 | 0.161 | -6.23 | 1.39 | 22% | 1% |
| Venular calibre uniformity | LV T2 mean (ms) | -0.14 | 0.594 | -0.14 | 0.08 | 54% | 0% |
| Venular calibre uniformity | PDAoD ($10^{-3}$mmHg$^{-1}$) | 0.10 | 0.462 | 0.09 | 0.04 | 48% | 0% |
| Arteriolar tortuosity | EAT (cm²) | 0.32 | 0.331 | 0.32 | 0.10 | 31% | 0% |
| Venular tortuosity | LAVi (ml/m²) | 1.93* | 0.042 | 2.01 | 0.38 | 19% | 57% |
| Venular tortuosity | LV T1 mean (ms) | 5.91 | 0.058 | 6.06 | 1.20 | 20% | 31% |
| Venular tortuosity | LV T2 mean (ms) | 0.46* | 0.013 | 0.47 | 0.09 | 18% | 89% |
| Venular tortuosity | LV GLS (%) | 0.63** | 0.006 | 0.64 | 0.09 | 14% | 93% |

Coefficient of variation of β: the standard deviation of the regression coefficient across the 100 re-sampled analyses divided by the mean coefficient (quantifying stability of the estimated effect size across the 100 re-samples)

% of β coefficients where p<0.05: the proportion of the 100 re-sampled analyses where the coefficient of the relationship between RVF and CMR was statistically significant (α=0.05)

*p<0.05 **p<0.01
